# Integration of CA attention and KAN algorithm to predict EGFR mutation status in lung cancer

**DOI:** 10.1101/2025.02.20.25322637

**Authors:** Nan Jia, Junfeng Xing, Siyuan Tang, Liang Liu, Jingxia Guo, Yan Wu, Jiniu Bai, Yan Li

## Abstract

Epidermal Growth Factor Receptor (EGFR) mutations are critical biomarkers for targeted therapies in non-small cell lung cancer (NSCLC). However, conventional diagnostic methods rely on invasive tissue biopsies, which are costly, time-consuming, and pose significant limitations. As an alternative, non-invasive approaches using lung CT imaging to predict EGFR mutation status have gained attention for their rapid and user-friendly nature. This study explores EGFR mutation prediction by analyzing seven distinct 2D regions of interest (ROI): the nodule itself, 2-pixel, 4-pixel, and 6-pixel extensions around the nodule, CT slices containing the nodule, single-lung segmented images with the nodule, and bilateral lung segmented images with the nodule. We developed a deep learning model combining EfficientNet-B0 with a Coordinate Attention (CA) mechanism, replacing the traditional MLP classifier with a KAN classifier. Results show that analyzing single-lung images containing the nodule captures additional relevant information, achieving the highest predictive performance. This suggests that regions surrounding the nodule contain valuable discriminatory biomarkers. The model achieved 92.73% accuracy on the single-lung test set with only 6.82M parameters, demonstrating its potential as a clinical tool to optimize targeted treatment decision-making.

## 1. Introduction

Cancer remains one of the leading causes of death worldwide. In 2022, there were 20 million new cancer cases and 9.7 million cancer-related deaths globally, with lung cancer being the most prevalent. Lung cancer alone accounted for 1.8 million deaths, representing 18.7% of all cancer-related fatalities and making it the leading cause of cancer mortality [1]. Non-small cell lung cancer (NSCLC) constitutes approximately 80% of all lung cancer cases, with adenocarcinoma being the most common histological subtype [2]. However, the efficacy of first-line chemotherapy for NSCLC is limited, with response rates of only about 30% [3]. Precision oncology, which tailors treatments based on a patient’s molecular profile, aims to improve therapeutic outcomes by identifying key genes involved in tumorigenesis. The advent of targeted therapies has revolutionized lung cancer treatment and improved clinical outcomes for certain patients. In regions such as China and other Asian countries, EGFR mutations are found in up to 30% of NSCLC cases, and epidermal growth factor receptor tyrosine kinase inhibitors (EGFR-TKIs) have demonstrated efficacy rates of up to 70% in these patients [44]. The 2021 NCCN guidelines (version 6) recommend molecular biomarker testing for NSCLC, including analysis of EGFR, ALK, KRAS, ROS1, BRAF, NTRK1/2/3, and METex14 skipping mutations. Among these, EGFR mutations are the most frequently observed genetic alterations in Chinese NSCLC patients.

Currently, molecular biomarker testing in clinical practice primarily relies on tissue biopsy. However, biopsies provide only a limited sample of tumor tissue, which may not fully capture tumor heterogeneity. Inadequate sampling or poor biopsy site selection can lead to false-negative results. Moreover, biopsies impose stringent requirements on patient conditions, tumor size, and location, and their associated costs limit widespread clinical use [5–7]. Computed tomography (CT) has emerged as a valuable tool for lung cancer screening, significantly reducing lung cancer mortality, and is widely employed in clinical settings. Nearly all lung cancer patients undergo multiple CT scans to monitor tumor progression. Extracting feature information from lung CT images holds significant potential for improving lung cancer screening, diagnosis, and treatment. In recent years, radiogenomics has gained considerable attention, with studies suggesting that CT features may be linked to gene expression [8, 9]. Establishing a connection between lung CT features and genetic mutations is therefore highly significant and holds broad potential applications.

In terms of image feature extraction, traditional radiomics and deep learning are the two primary methods currently in use. Traditional radiomics involves transforming images into high-dimensional, analyzable data for subsequent analysis, facilitating improved decision support. Common radiomics features include assessments of size, shape, and texture, and have been applied across imaging of various organs and systems, such as the brain, pituitary, lungs, heart, liver, kidneys, adrenal glands, and prostate. In recent years, deep learning has emerged as a dominant approach. This technique requires only pre-processed data and utilizes self-learning algorithms to automatically extract image features. Deep learning has demonstrated exceptional diagnostic capabilities in fields such as retinal diseases, skin cancer, and breast cancer, often surpassing the diagnostic accuracy of experienced clinicians [10, 11].

## 2. Related work

Radiomics typically involves several key steps: image acquisition, ROI segmentation, feature extraction, feature selection, and classifier model development [12]. Extensive research has been conducted on the use of radiomics in predicting genetic mutations. For example, Le et al. [13] utilized the NSCLC-Radiogenomics public dataset to extract features from lung nodules using conventional radiomics techniques. They employed genetic algorithms and the XGBoost model to predict EGFR and KRAS mutations, achieving accuracies of 0.836 and 0.86, respectively. Similarly, Pinheiro et al. [14] explored the relationship between lung CT imaging and EGFR/KRAS mutations using the same dataset. They combined imaging features, clinical data, and semantic features of lung nodules, inputting these into an XGBoost model. Their results showed that including all features yielded the best performance, with an AUC of 0.746 ± 0.088 for EGFR mutations and 0.504 ± 0.078 for KRAS mutations. Liu et al. [15] developed a multi-center lung CT dataset, using data from three centers for training and one for validation. They manually annotated lesion ROIs and applied conventional radiomics methods for feature extraction. Four machine learning models (LR, DT, RF, and SVM) were trained, with the Random Forest (RF) model outperforming the others. Similarly, Hong et al. [16] used radiomics to extract features and trained six machine learning models (LR, DT, RF, SVM, NBC, and KNN). Their results indicated that logistic regression (LR) achieved the best performance, with an AUC of 0.851 on the validation set. Nair et al. [17] extracted radiomic features from CT and PET-CT images to predict EGFR mutations in NSCLC. Their analysis revealed that PET-CT-derived features were more effective than CT features in identifying EGFR mutations and distinguishing between exon 19 and exon 21 mutations. Zhang et al. [18] applied radiomics to identify multiple gene mutations (EGFR, KRAS, ERBB2, and TP53) in NSCLC. By integrating radiomic and clinical features, they trained a logistic regression model, achieving AUC values of 0.78, 0.81, 0.87, and 0.84 for the respective mutations.

Some researchers have expanded ROI selection to include the entire lung or both lungs containing the nodule, aiming to reveal genetic mutation-related information beyond the nodule itself. Morgado et al. [19] utilized the NSCLC-Radiogenomics dataset, selecting the affected lung as the ROI. They applied conventional 2radiomics methods for feature extraction and selection, followed by machine learning-based prediction. Their results showed that linear support vector machines, elastic net models, and logistic regression, combined with PCA for feature selection, achieved the best performance, with AUC values ranging from 0.725 to 0.737. Chen et al. [20] constructed a private CT dataset of 233 lung cancer cases with EGFR mutations. Using a semi-automatic approach to delineate the affected lung regions, they incorporated clinical data into a logistic regression model to predict EGFR mutation types and subtypes (exon 19 deletion and exon 21 L858R mutation). Their results demonstrated an AUC of 0.759 for predicting mutation types and 0.554 for subtypes.

With the advancement of deep learning, researchers have adopted holistic and end-to-end approaches to explore the correlation between imaging features and genetic mutations, offering higher accuracy and robustness. Typically, two methods are used for deep learning-based mutation prediction. The first involves constructing a 2D deep learning model, where clinicians manually annotate tumor regions and feed the ROI-cropped images into the model. For instance, Huang et al. [5] collected CT images from 228 lung adenocarcinoma patients (116 EGFR mutant and 112 wild-type). Due to the small sample size, they used all slice images of each lesion for training a ResNet model, achieving higher prediction accuracy compared to traditional radiomics. Wang et al. [21] collected preoperative CT images and clinical data from 844 lung adenocarcinoma patients across two hospitals. They employed the DenseNet model to predict EGFR mutations from 14,926 CT images, achieving an AUC of 0.85 on the training set and 0.81 on the test set.The second method involves 3D deep learning models. For example, Wang et al. [23] proposed a 3D CNN model to classify EGFR and PDL1 mutations using lung CT images, achieving an AUC of 0.96 on the training set and 0.76 on the validation and test sets. Zhao et al. [24] developed a 3D DenseNets system to automatically predict EGFR-mutant lung adenocarcinoma, applying the Mixup data augmentation technique to improve generalization. Their model achieved AUC values of 75.8% on a private test set and 75.0% on a public test set.

Despite these advancements, most studies still rely on manual annotation of lung and lesion regions by clinicians, which is time-consuming and expertise-dependent. Automating ROI annotation is crucial to enhance data labeling efficiency and facilitate the deployment of automated gene prediction systems. Current approaches primarily use 2D or 3D models, such as ResNet, DenseNet, or 3D CNN variants, to process CT images.

The primary contributions of this study are as follows:

1. We processed a publicly available lung nodule CT dataset to create seven distinct datasets, including 2D nodule images, expanded nodule regions, and full lung datasets.

2. We employed a “human-machine collaborative” approach for lung contour segmentation, significantly improving annotation efficiency.

3. We designed an improved EfficientNet-B0 architecture with a CA attention mechanism and a KAN classification head, enhancing feature extraction and classification capabilities. Experimental results demonstrate that the proposed model outperforms existing approaches.

## 3. Dataset and Preprocessing

### Dataset

This study employs two publicly available datasets. The first dataset, COVID-19-CT-Seg_20cases [27], comprises 20 labeled COVID-19 CT scans. The left lung, right lung, and infection areas were annotated by two radiologists and subsequently validated by an experienced radiologist. In this study, only the original CT images and the annotations of the left and right lungs were utilized.

The second dataset is the NSCLC Radiogenomics dataset [28], which contains chest CT scans, tumor segmentation masks, and EGFR mutation status for non-small cell lung cancer (NSCLC) patients. This dataset includes 211 retrospectively collected cases from the Stanford University School of Medicine and the Palo Alto Veterans Affairs Health Care System. The CT scans were acquired using various scanner models and protocols, with slice thicknesses ranging from 0.625 to 3.0 mm (median: 1.5 mm). The X-ray tube currents varied between 124 and 699 mA (mean: 220 mA), and the tube potentials ranged from 80 to 140 kVp (mean: 120 kVp). After data review, patients were selected 3based on the availability of tumor binary masks and EGFR mutation results. A total of 116 patients met the inclusion criteria, including 23 with EGFR mutations and 93 with wild-type EGFR (see Table 1).

**Table 1:**
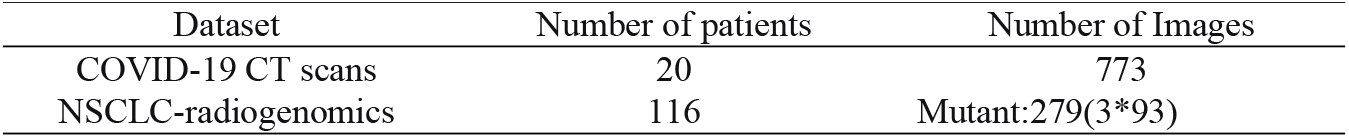
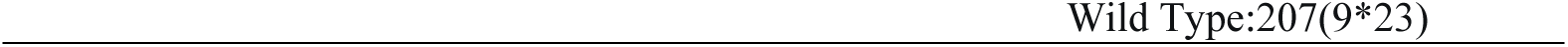
Summary of the number of images used for each task.

| Dataset | Number of patients | Number of Images |
| --- | --- | --- |
| COVID-19 CT scans | 20 | 773 |
| NSCLC-radiogenomics | 116 | Mutant:279(3*93) |

### Preprocessing

The COVID-19 CT scans dataset is stored in NIFTY format, whereas the NSCLC Radiogenomics dataset is in DICOM format. To ensure consistency, we first converted the DICOM images from the NSCLC Radiogenomics dataset into NIFTY format. Subsequently, all CT images and their corresponding masks from both datasets were resampled to a uniform voxel size of 1 mm × 1 mm × 1 mm. Additionally, the CT images were adjusted to a Hounsfield Units (HU) range of [-1000, 400] and normalized to the range [0, 1].

For the COVID-19 CT scans dataset, we focused on lung CT images and their associated lung masks. From each case, we extracted all images with corresponding lung masks, resulting in a total of 2,973 2D images. Figure 1 illustrates a sample of the extracted 2D images and their corresponding lung masks.

For the NSCLC Radiogenomics dataset, we followed a previous study [29] and randomly divided the dataset into training and testing subsets at an 80:20 ratio. To address class imbalance and facilitate model training, we adopted a selective sampling strategy. For mutant cases (23 samples), we selected the axial slice with the largest tumor area, along with the four preceding and subsequent slices, yielding a total of 9 2D images per case. For wild-type cases (93 samples), we selected the axial slice with the largest tumor area and its immediate adjacent slices, resulting in 3 2D images per case. This approach helped balance the dataset across different image types. Figure 2 displays examples of the processed 2D images.

**Figure 1:**
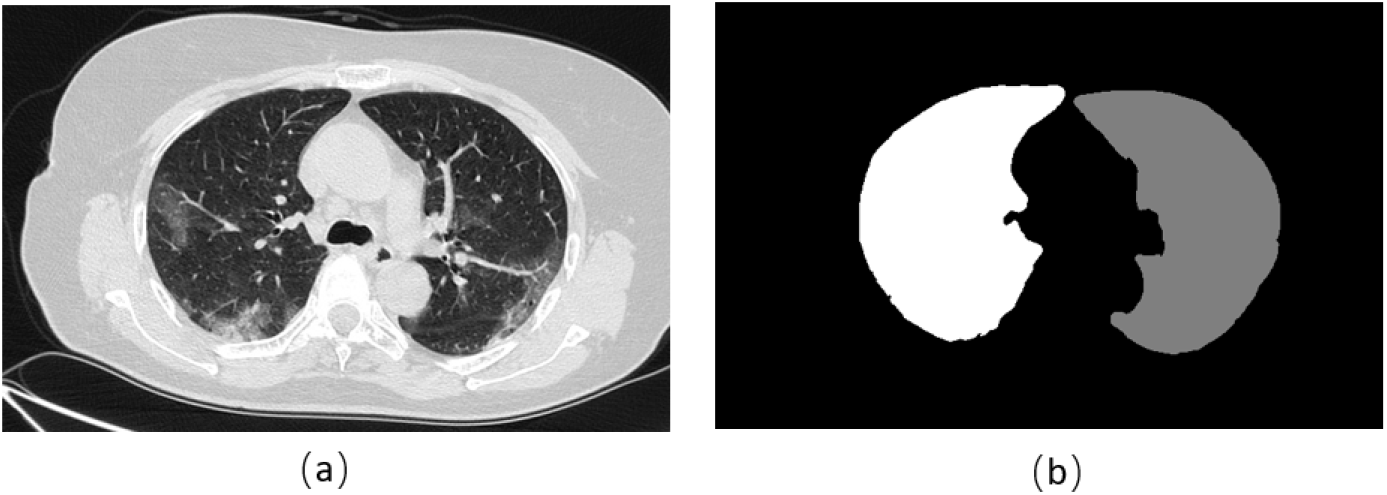
Lung image slices (a) and left and right lung masks (b).

**Figure 2:**
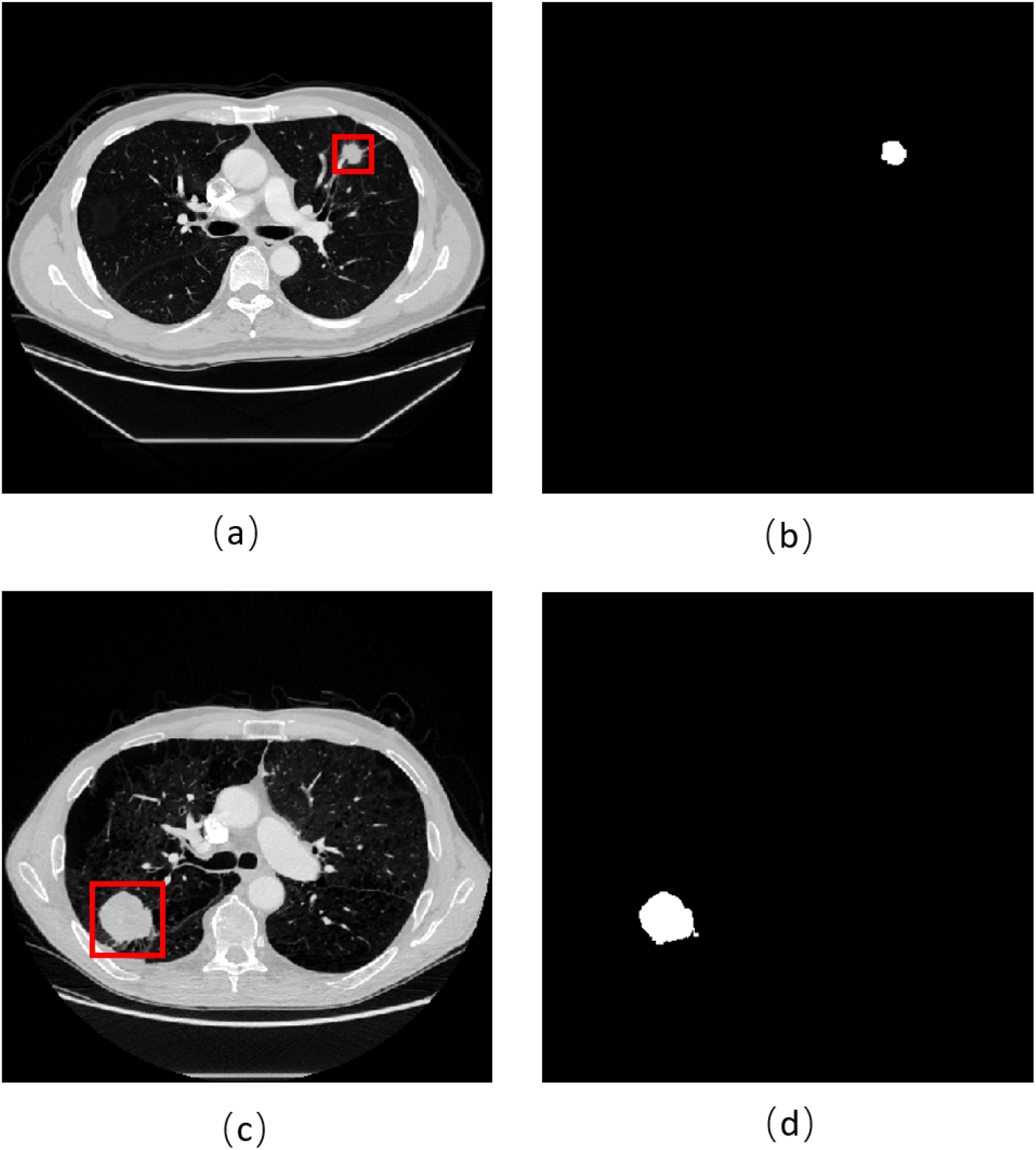
Mutant-type EGFR image and mask are shown in (a) and (b). Wild-type EGFR image and mask are shown in (c) and (d).

## 4. Methods

The methodology of this study comprises two main stages. The first stage involves the construction of seven 2D image datasets, while the second stage focuses on developing an end-to-end classification model for predicting EGFR mutation status. This model integrates an EfficientNet_CA encoder with a KAN classifier. An overview of the methodology is illustrated in Figure 3.

**Figure 3:**
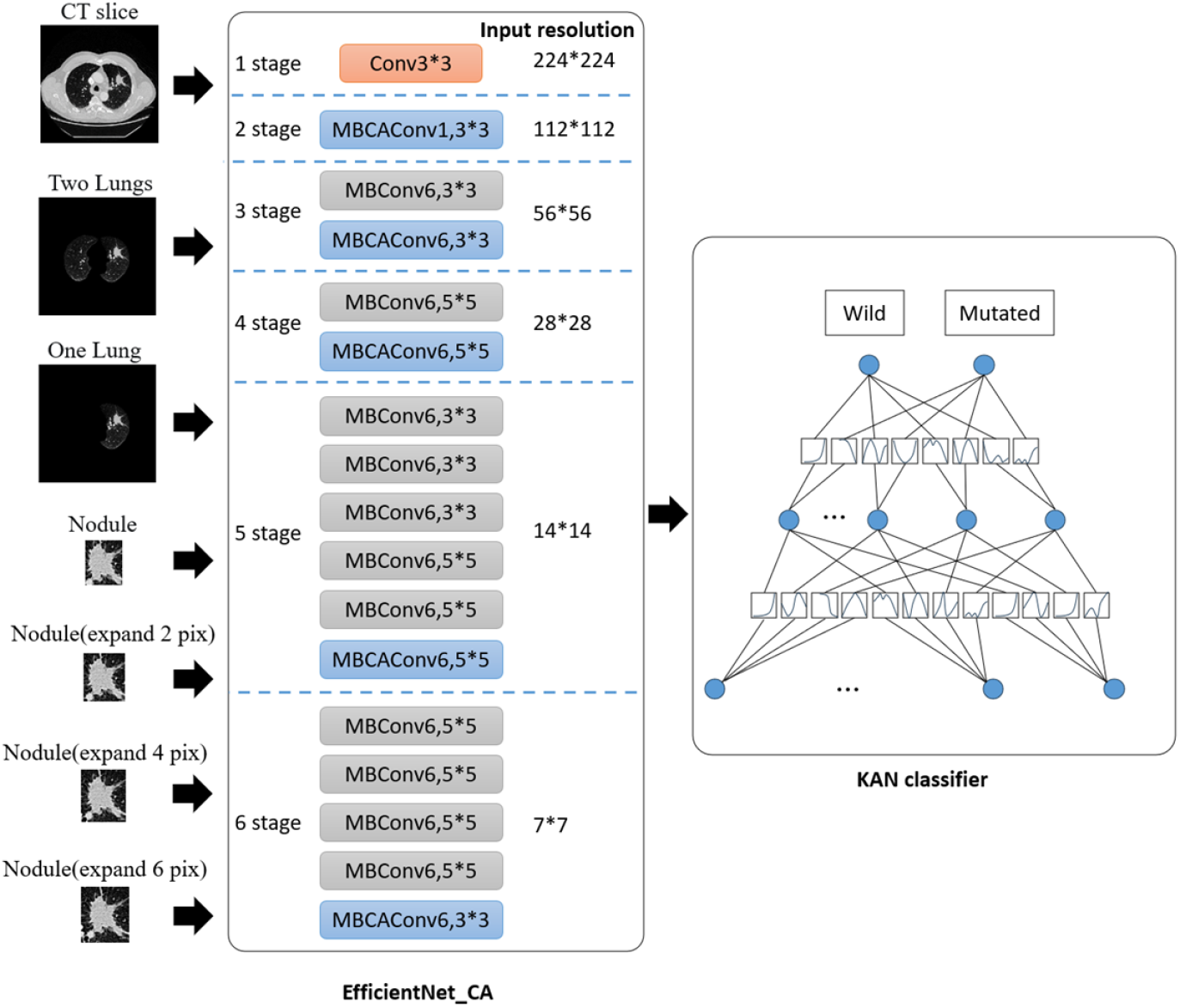
Overview of the proposed approach

### The “human-computer collaboration” annotation scheme for lung contour segmentation

If the model is trained directly on CT slices containing lung nodules, the prediction accuracy is notably high. However, upon analyzing the heatmap (as depicted in Figure 7), we observed that the model’s classification of gene mutation status did not primarily rely on information from the lung nodules or lung regions. Instead, the model focused on areas outside the lungs, suggesting that it failed to correctly identify the relevant regions of interest in the images. Consequently, the predictions of gene mutations lacked interpretability.

To address this issue, we first extracted the bilateral lung regions from the dataset. Subsequently, we constructed a training dataset comprising images with lung nodules from either a single lung or both lungs. Previous studies often required experienced physicians to manually annotate lung regions from raw data using software such as 3D Slicer or ITK-SNAP, a process that is both time-consuming and labor-intensive.

In this study, we adopted a more efficient approach. We initially trained a bilateral lung segmentation model (KNet) [30] using a publicly available COVID-19 CT scans dataset with lung masks. The 2D image data from the target dataset were then input into this model to generate corresponding bilateral lung segmentation masks. The resulting masks were saved in PNG format, and multiple consecutive PNG mask images for a single patient were consolidated into a single NIFTY file. Physicians reviewed the model’s predictions using ITK-SNAP, made necessary adjustments, and saved the modified files. Finally, the revised NIFTY files were converted back into individual image slices.

The overall workflow is illustrated in Figure 4. We found that this approach generally produced highly accurate lung contour masks. In rare cases, particularly near the lung boundaries, the model generated incorrect predictions. However, these errors could be easily corrected by physicians with minor adjustments, significantly improving labeling efficiency. Moreover, the quality of the annotations achieved through this semi-automated process was superior to that of fully manual annotation.

**Figure 4:**
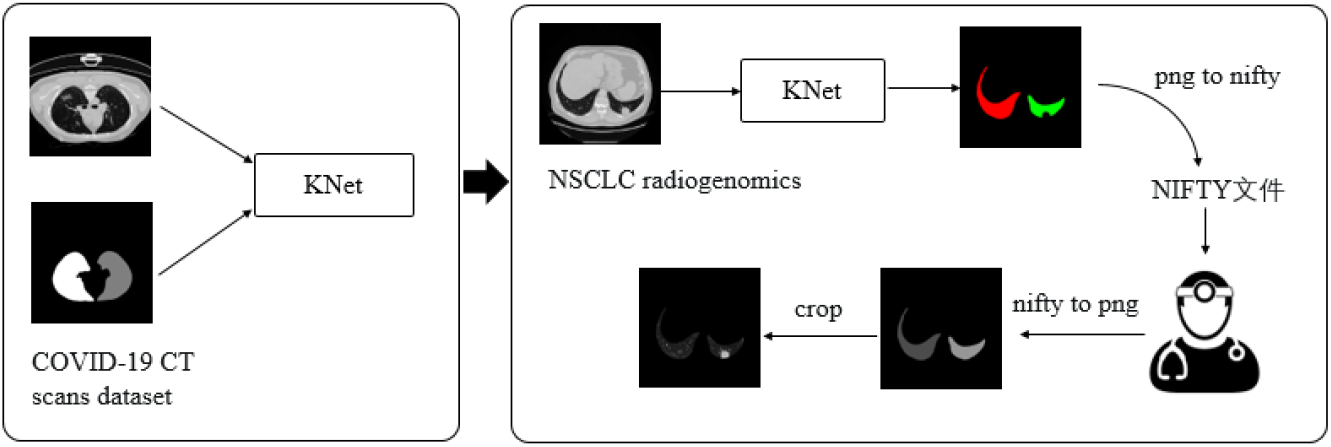
Human-AI Collaborative Annotation Pipeline for Accurate Lung Contour Segmentation in Medical Imaging

### EfficientNet_CA Network

EfficientNet [31], introduced by the Google Research team in 2019, is a highly efficient convolutional neural network (CNN) model. Its core design emphasizes balancing the network’s depth, width, and input resolution to achieve both computational efficiency and superior performance. Through the innovative use of compound scaling, EfficientNet optimizes its architecture, significantly reducing the number of parameters and computational costs while maintaining high accuracy.

In this paper, we propose EfficientNet_CA, an enhanced version of EfficientNet_B0. As illustrated in Figure 3, the network is divided into six stages, with the feature map size halving at each stage. Starting from an initial size of 224×224 at Stage 1, the feature map reduces to 7×7 by Stage 6. Concurrently, the number of channels in the feature maps increases progressively from Stage 2, culminating in 1280 channels at the output of Stage 6.

The EfficientNet_CA network primarily comprises two modules: MBConv and MBCAConv. The MBConv module, inherited from the original EfficientNet, is depicted in Figure 5. In contrast, the MBCAConv module replaces the Squeeze-and-Excitation (SE) attention mechanism in MBConv with the CA mechanism [32], as shown in Figure 6. The CA mechanism is designed to improve a model’s ability to capture spatial relationships within input data. Its key innovation lies in integrating coordinate information, enabling the model to better understand positional dependencies. Specifically, the CA mechanism applies global average pooling twice to the input feature map—once along the width and once along the height. These operations generate two feature maps, z^w^ and z^h^, which encode global features in the width and height directions, respectively, as described by the following equations:

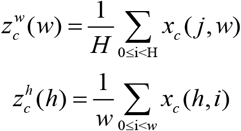

The two feature maps described earlier are merged, and a simple concatenation operation is applied to form a new feature layer with dimensions [C, 1, W+H], where W+H represents the combined width and height dimensions. A convolution operation is then performed on the merged feature layer with a 1×1 kernel, reducing the output dimensions to C/r. Following batch normalization, a nonlinear activation function is applied to the feature map F1, resulting in a feature layer output of shape 1 × (W+H) × C/r, where *f* ∈ *R*^*C*/*r*×(*H* +*W*)^, as shown in the following equation:

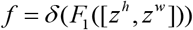

Subsequently, the feature map f is split along the spatial dimensions into two distinct tensors, f^h^, and f^w^. Two 1×1 convolutions are then applied to transform these feature maps (f^h^ and f^w^) to match the same number of channels as the input x.

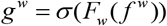

Next, the tensors g^h^ and g^w^ are expanded and used as attention weights, which are then multiplied by the input. The final output of the CA module is represented by the following equation.

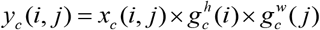

In the EfficientNet_CA architecture, commencing from the second stage, the concluding module of each stage is designated as MBCAConv, while the remaining modules are consistently MBConv.

**Figure 5:**
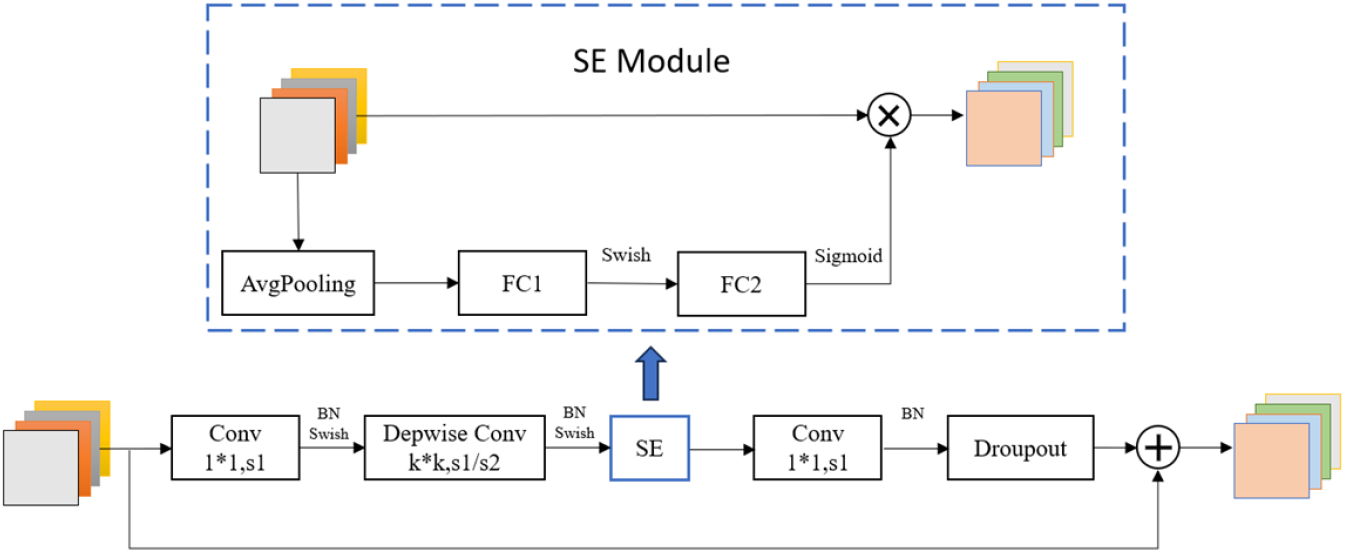
MBConv Architecture Diagram, which distinctly delineates the constituent elements of MBConv and the trajectory of data flow within the architecture.

**Figure 6:**
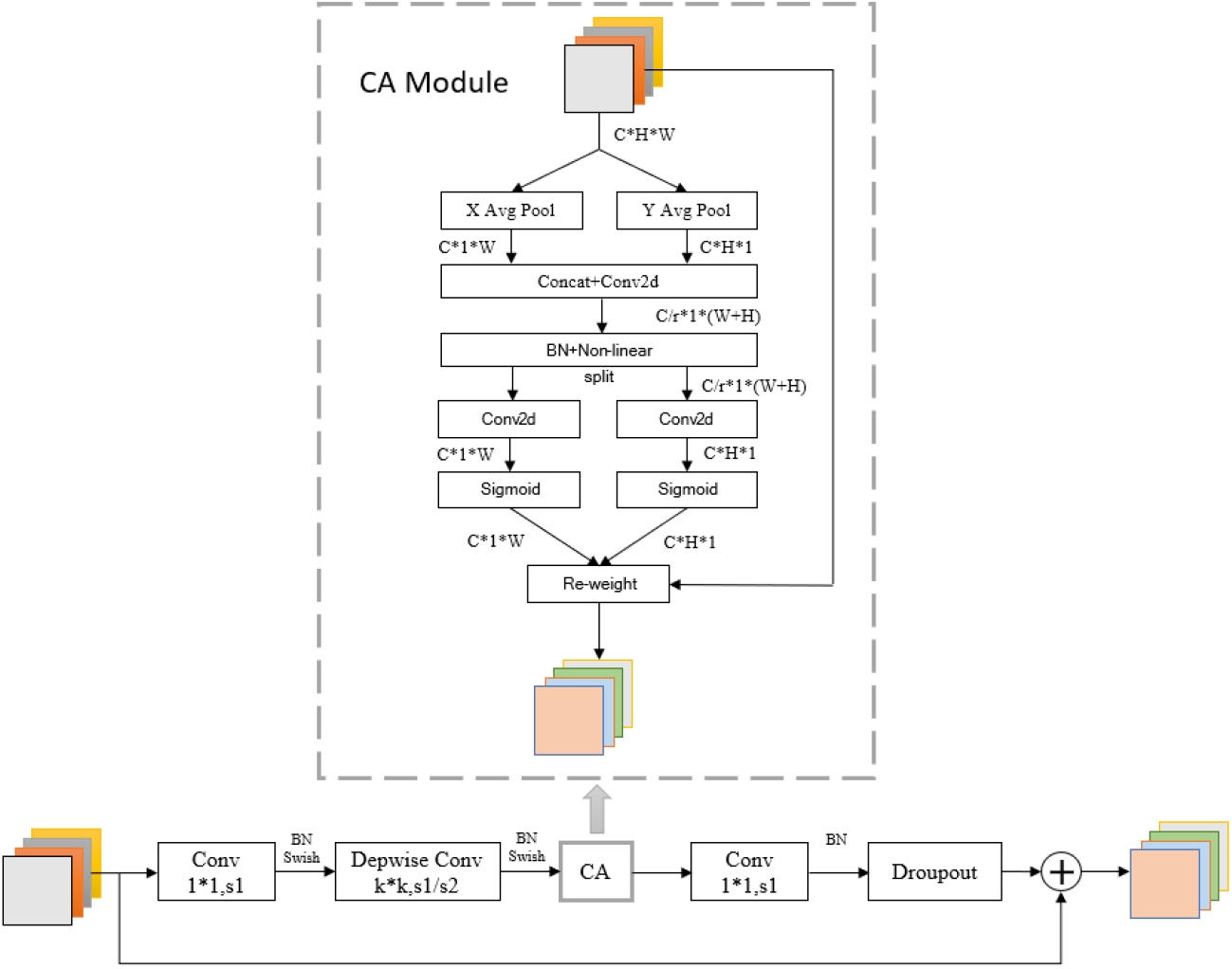
MBCAConv Architecture Diagram, which distinctly delineates the constituent elements of MBCAConv and the trajectory of data flow within the architecture.

### KAN Classification Head

KAN [33] offers a promising alternative to traditional Multilayer Perceptrons (MLPs). Unlike MLPs, which rely on the universal approximation theorem and employ fixed activation functions at nodes (neurons), KAN is grounded in the Kolmogorov-Arnold representation theorem. In KAN, activation functions are applied at the edges (weights) and are learnable.

Additionally, KAN does not use linear weights; instead, each weight parameter is represented as a spline unary function. The computation of an L-layer KAN is defined as follows: KAN(x) = (Φ _L-1_ ° Φ _L-2_ °‥.Φ °^1^ ° Φ _0_)(X), where Φ _L-1_ is the function matrix of the (L-1)-th layer.

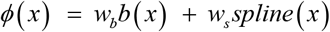

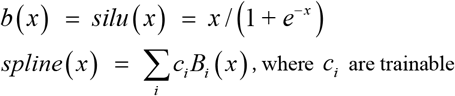

By leveraging the expressive power of spline functions, KAN achieves strong representational capabilities with fewer nodes, significantly reducing the number of parameters compared to traditional MLPs.

In this study, after extracting image features using the EfficientNet_CA network, we replaced the conventional MLP with a KAN network for classification. The architecture of the KAN network is depicted in Figure 3. The number of input nodes matches the output feature channels of the EfficientNet_CA network (1280 channels). The hidden layer consists of 16 nodes, while the output layer comprises 2 nodes, corresponding to the two possible EGFR mutation statuses.

## 5. Results

### Implement Detail

All models in this experiment were trained on an NVIDIA GeForce RTX 3090 graphics card running the Linux operating system. The deep learning framework used for training was PyTorch 2.1.1.

The learning rate is determined using the Multiple learning rate schedule, where the first 20 epochs utilize linear warm-up by epoch, followed by a CosineAnnealing schedule. The optimizer used is AdamW, with weight_decay set to 0.3. The learning rate is initialized to 0.0005. The entire training process is halted at the 400th epoch.

### EGFR mutation status classification

We conducted seven experiments to evaluate different ROI selections in the lungs, including CT slices containing nodules, bilateral lungs with nodules, single lung with nodules, the nodule itself, and nodule regions extended by 2, 4, and 6 pixels. The dataset was randomly split into training and testing sets in an 80:20 ratio, repeated five times. The results for each ROI on the test set are presented in Table 2.

**Table 2:**
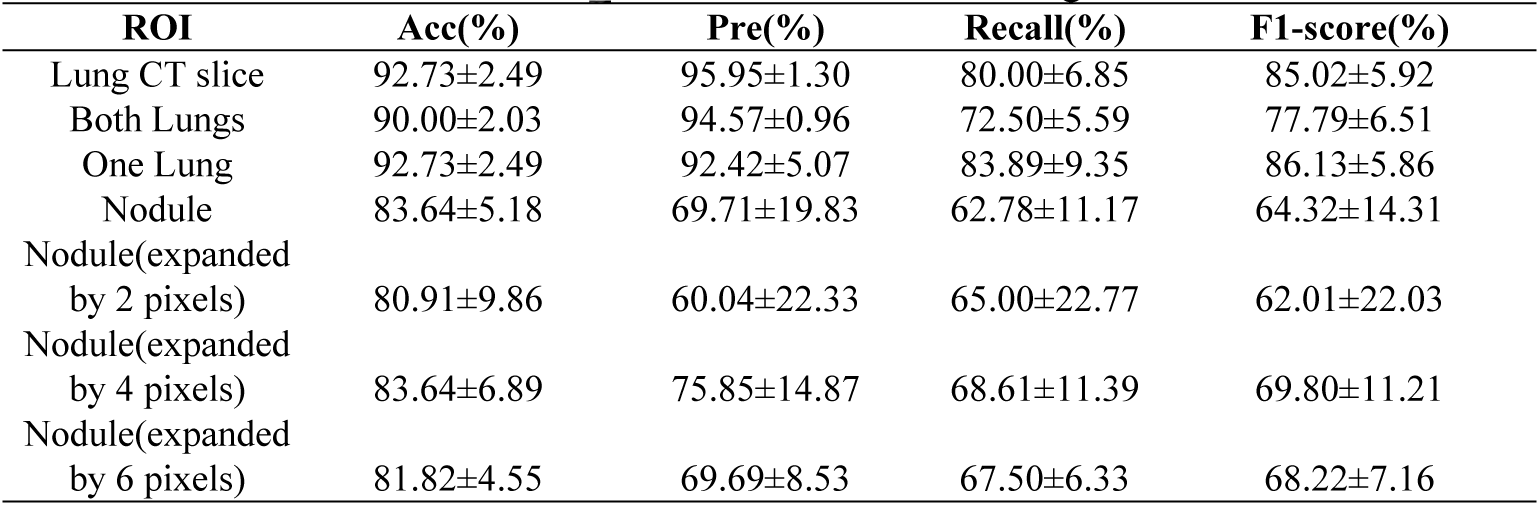
Performance of EfficientNet_CA across different ROI regions.

| ROI | Acc(%) | Pre(%) | Recall(%) | F1-score(%) |
| --- | --- | --- | --- | --- |
| Lung CT slice | 92.73±2.49 | 95.95±1.30 | 80.00±6.85 | 85.02±5.92 |
| Both Lungs | 90.00±2.03 | 94.57±0.96 | 72.50±5.59 | 77.79±6.51 |
| One Lung | 92.73±2.49 | 92.42±5.07 | 83.89±9.35 | 86.13±5.86 |
| Nodule | 83.64±5.18 | 69.71±19.83 | 62.78±11.17 | 64.32±14.31 |
| Nodule(expanded<br>by 2 pixels) | 80.91±9.86 | 60.04±22.33 | 65.00±22.77 | 62.01±22.03 |
| Nodule(expanded<br>by 4 pixels) | 83.64±6.89 | 75.85±14.87 | 68.61±11.39 | 69.80±11.21 |
| Nodule(expanded<br>by 6 pixels) | 81.82±4.55 | 69.69±8.53 | 67.50±6.33 | 68.22±7.16 |

The results in Table 2 show significant variance in the model’s performance metrics, likely due to the limited dataset size, which includes only 23 patients with EGFR mutations. When the ROI is limited to the lung nodule, the model achieves an accuracy of 83.64% (Table 2). However, expanding the nodule region by 2, 4, or 6 pixels does not improve performance. For instance, expanding the nodule by 6 pixels reduces accuracy to 81.82%, indicating that including the surrounding nodule area in the ROI does not enhance feature extraction for EGFR mutation prediction. These results suggest that the surrounding nodule area does not provide additional predictive value for EGFR mutation status.

Expanding the ROI to include the entire single lung containing the nodule significantly improves performance, achieving an accuracy of 92.73%. In contrast, extending the ROI to both lungs slightly reduces accuracy to 90.00%. The model also demonstrates strong performance when the ROI consists of a CT slice containing the entire tumor, maintaining an accuracy of 92.73%.

However, analysis of spatial heatmaps (Figure 7) reveals that the model’s decisions for EGFR mutation status are not primarily based on nodule or surrounding lung features, as evidenced by four randomly selected test images (R01-050, R01-066, R01-076, R01-099). This suggests that relying solely on CT slice images may be suboptimal. This suggests that relying solely on CT slice images for modeling may be problematic. Therefore, modeling based on single-lung images containing the nodule is the most effective approach, achieving the highest accuracy.

**Table 3:** Comparative performance of different methods.

| Method | Bankbone | Acc(%) | Pre(%) | Rec(%) | F1(%) | Flops(G) | Params(M) |
| --- | --- | --- | --- | --- | --- | --- | --- |
| VGG | VGG16 | 89.09±5.18 | 81.89±23.52 | 73.89±18.09 | 74.08±18.87 | 15.46G | 130M |
| EfficientNet | b0 | 90.91±3.21 | 90.93±5.89 | 78.89±10.32 | 81.56±9.45 | 0.4G | 6.75M |
|  | b3 | 90.00±4.98 | 85.86±11.79 | 78.33±11.44 | 80.61±11.42 | 0.99G | 10.70M |
| MobileNetV2 |  | 90.91±5.57 | 95.14±2.92 | 75.00±15.31 | 78.74±13.93 | 0.313G | 6.68M |
| Restnet | Restnet50 | 90.00±3.80 | 90.19±7.08 | 76.39±12.73 | 79.02±9.84 | 4.11G | 23.51M |
|  | Restnet101 | 90.91±4.55 | 95.09±2.26 | 75.00±12.50 | 79.20±12.71 | 7.83G | 42.50M |
| Vision Transformer | vit-base-p16 | 88.18±2.49 | 86.65±6.29 | 73.33±11.13 | 75.25±9.29 | 16.78G | 85.80M |
| Swin-Transformer | Swin-b | 91.82±4.98 | 93.52±5.57 | 79.44±13.94 | 82.45±12.09 | 15.47G | 86.75M |
| **Ours3** |  | 92.73±2.49 | 92.42±5.07 | 83.89±9.35 | 86.13±5.86 | 0.404G | 6.82M |

**Figure 7:**
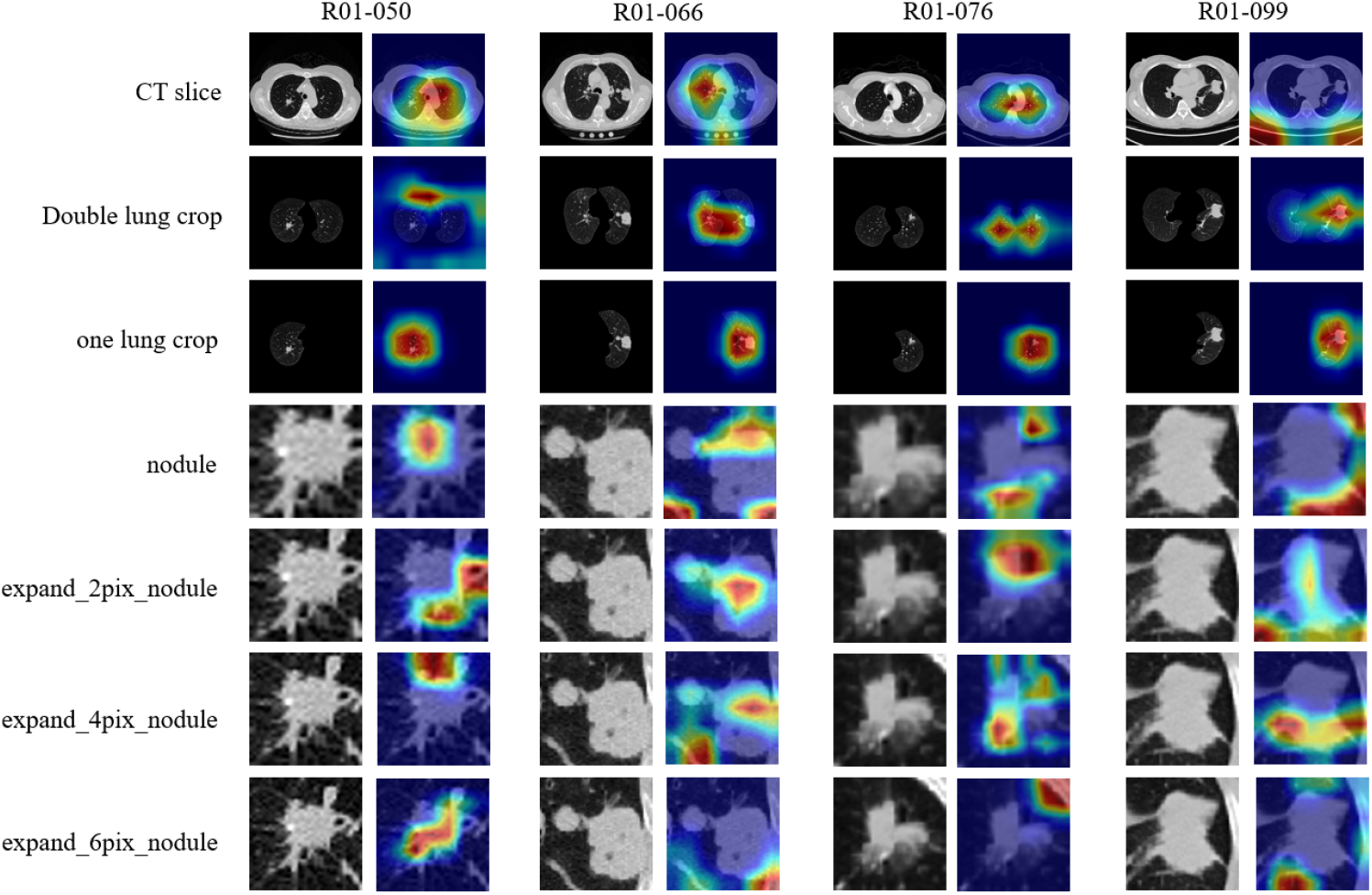
Visualization of Heatmaps in EfficientNet_CA for Seven Different ROI Datasets. The heatmaps, generated using the gradient-based XGrad-CAM method, demonstrate the model’s attention distribution across various ROI, highlighting the areas that contribute most to the prediction of EGFR mutation status.

## 6. Discussion

The proposed EfficientNet-CA model achieves state-of-the-art (SOTA) performance in predicting EGFR mutation status from single-lung segmented images containing lung nodules. The model achieves an accuracy of 92.73%, surpassing the second-place Swin-Transformer by 0.91%. With only 6.82M parameters, the model is comparable to EfficientNet-B0 (6.75M parameters), demonstrating high efficiency and suitability for mobile deployment.

This research also reaffirms the relationship between imaging features of the tumor’s surrounding region and genomic data. However, selecting the optimal surrounding region remains challenging. Our experiments show that extending the tumor by 2, 4, or 6 pixels significantly underperforms compared to single-lung segmentation containing the nodule. A potential reason is that small lung tumors may not provide sufficient discriminative information when extended by a few pixels, limiting the model’s ability to extract relevant features for classification. Additionally, as seen in the heatmap for R01-076 with a 6-pixel extension (Figure 7), including parts of bone structures in the tumor extension can greatly affect the model’s ability to discriminate and, consequently, degrade the final results.

In future work, we aim to explore the following areas. First, we will investigate the integration of state-space models (such as Mamba) with Vision Transformers (ViT) or Convolutional Neural Networks (CNNs) to further improve classification accuracy. Second, we plan to collect data from a broader range of sources and create a multi-center dataset to better evaluate the model’s generalizability. Lastly, we will focus on standardizing the analysis of the tumor surrounding region and integrating multimodal data to enhance predictive performance.

## 7. Conclusion

This study presents a novel method that combines EfficientNet-B0 with the CA attention mechanism as a feature extractor, followed by the use of KAN as a replacement for the traditional MLP in the final classifier for EGFR mutation status prediction. In this work, we thoroughly analyze the influence of images from different lung regions on the classification outcomes for EGFR mutation status. The results indicate that images from the single-lung region containing the nodule outperform other ROI regions, as they more effectively capture features relevant to lung cancer. Compared to other recognition methods, our proposed approach demonstrates superior classification accuracy, reduced model complexity, and provides valuable support for clinical diagnosis. It is also easy to deploy and holds significant potential for clinical application.

## Data availability

The two public datasets used in this study are available in the following sites:

(1) COVID-19-CT-Seg_20cases: https://zenodo.org/records/3757476;

(2) NSCLC radiogenomics: https://www.cancerimagingarchive.net/collection/nsclc-radiogenomics/;

Upon the publication of the research paper, the seven 2D datasets derived from the processing of the publicly available NSCLC radiogenomics dataset, along with the source code utilized in this study, will be made publicly accessible on the GitHub platform.

## Notes

### Competing Interest Statement

The authors have declared no competing interest.

### Funding Statement

Yes

### Author Declarations

The datasets used in this paper are all publicly available datasets.

